# Interictal epileptiform discharges are involved in momentary lapses of attention in children with epilepsy

**DOI:** 10.1101/2025.07.11.25331382

**Authors:** Marine Thieux, Romain Bouet, Julien Jung, Philippe Kahane, Laurent Vercueil, Lucie Martinet, Marcela Perrone-Bertolotti, Jean-Philippe Lachaux, Julitta de Bellescize, Vania Herbillon

**Author notes:** **Corresponding author:** Marine Thieux.

## Abstract

**Objective:** Attention impairments are common in children with epilepsy and widely impact their quality of life. Interictal epileptiform discharges (IED) may induce subtle dysfunctions of various cognitive processes, but data regarding the impact of IED on attention remain limited. The objective of the present study was to evaluate the impact of IED on continuous undivided attention in children with epilepsy, controlling for the number of treatments, type of epilepsy, frequency of seizures in the last year, age at onset, and comorbid attention disorder.

**Methods:** Using a computerized attention test synchronized with the electroencephalogram (EEG) in 119 children with diverse epilepsy syndromes, behavioral (reaction time [RT], errors, attention stability over time) and electrophysiological measures related to attentional engagement (event-related potential [ERP] in parietal electrodes) were collected. The cumulative impact of IED was evaluated using multivariate models controlling for epilepsy-related factors. The transient impact of IED was assessed by comparing responses in trials with and without IED.

**Results:** IED induced attention fluctuations independently from other epilepsy-related factors. In terms of cumulative impact, a higher quantity of IED was associated with a poorer attentional performance over the entire task. In terms of transient impact, trials disrupted by IED were characterized by longer RT and a lower amplitude of the parietal ERP, extending over a long-time window that included attentional processing (P300).

**Significance:** These results highlight the deleterious effect of IED on attention, at both behavioral and electrophysiological levels, independently of other epilepsy-related features. This work provides evidence that IED induce subtle attentional deficits and could be considered as biomarkers of abnormal brain function.

**Key points:**

- A higher quantity of IED was associated with poorer attentional performance, irrespective of other epilepsy-related features.
- IED promote transient cognitive impairment: trials with IED are characterized by increased RTs and a lower amplitude of the parietal ERP.
- EEG-synchronized attentional tasks are needed to assess subtle deficits in patients with epilepsy.

## 1. Introduction

Attention impairments are particularly common in children with epilepsy and widely impact their quality of life.^1^ Although generally attributed to epilepsy-related features (*e.g.* seizures, lesions, treatments),^2^ these impairments can persist in patients whose epilepsy is well stabilized and in the absence of brain damage.^3^ The seminal work of Aarts *et al.*, reported that interictal epileptiform discharges (IED) were concomitant with transient memory impairment. Various data gathered during the last forty years have also suggested that IED may induce subtle dysfunctions of various cognitive processes;^4–10^ these studies mostly used electroencephalogram (EEG) along with neuropsychological testing. Furthermore, recent studies using intracranial EEG and trial-by-trial analyses captured IED-induced disturbances in attentional set-shifting and word-finding.^12,13^ Nevertheless, data regarding the impact of IED on attention impairments remain limited, notably due to assessment methods. Traditional attention tests are well-designed to detect long-lasting, slow drifts away from the ongoing task. Brief lapses of attention may therefore go unnoticed, particularly when only a subset of stimuli requires attention, or when performance is averaged on the entire task.^11^

The objective of the present study was to evaluate the impact of IED on continuous undivided attention in children with epilepsy. The Bron-Lyon Attention Stability Test (BLAST) is specifically designed to capture brief lapses of attention by combining trial-by-trial measures (*i.e.* accuracy and speed) with variables reflecting the dynamics of attention engagement over the task (*i.e.* series of error-free responses with fast or steady response times). It has previously been used to assess the ability to stay-on-task on a second-by-second basis.^11,14^ Therefore, the concomitant use of the BLAST and the EEG would allow to: (i) assess the cumulative IED impact at the behavioral level by controlling for other epilepsy-related features (*i.e.* number of anti-seizure medication, type of epilepsy, frequency of seizures in the last year, age of onset, and comorbid attention disorder); and (ii) assess the transient IED impact at the behavioral level as well as at the electrophysiological level by a modulation of the parietal eventrelated potential (ERP), with a primary focus on a P300 like, which is known to reflect attentional engagement.^15^

## 2. Materials and methods

### 2.1. Patients

A total of 119 children (from 6 to 18 years old) with epilepsy who underwent a 30-minute EEG recording for clinical follow-up were included in this study between 2015 and 2017. The inclusion criteria were: (i) regular school curriculum; (ii) absence of major visual, auditory, or upper limb motor impairment; and (iii) absence of epileptic seizure during the concomitant BLAST-EEG or in the few hours before. Children with epilepsy were included irrespective of syndrome, seizure type (focal or generalized) and frequency over the last 12 months, or number of antiseizure medication (ASM). Diagnosis was confirmed by experienced neurologists (J.D.B.) according to the International League Against Epilepsy criteria.^16^ Presence of a comorbid attention deficit hyperactivity disorder (ADHD) was assessed by an experienced neuropsychologist (V.H.) using the ADHD rating scale (ADHD-RS) and the Diagnostic and Statistical Manual of Mental Disorders (DSM-V).^17,18^ No children with epilepsy was treated with a psychostimulant. The study was approved by an institutional review board (COGNIT-AIC-38RC14.374) and informed written consent was obtained from each child and their care-givers. The study was registered on ClinicalTrials.gov (NCT03094793). Some of the data had already been published in a previous study.^11^

### 2.2. Procedure

#### 2.2.1. Cognitive task

The BLAST has been validated in a large cohort of healthy volunteers.^11,14^ The present study is part of a broader research that use all versions of BLAST. Herein, a child-friendly version is employed. In each trial, a letter (randomly chosen among 12) appeared on the screen (in foveal vision), then was replaced by a mask (#) and then, by a 2-by-2 array of 4 letters (randomly selected from the alphabet) in which one of them was red half the time (**Figure 1**). Children with epilepsy had to determine the presence of a red letter, irrespective of the initial letter, by answering “yes” with their non-dominant hand or “no” with their dominant hand. They were told they must find a balance between speed and accuracy. After a few practice trials, 3 BLAST sessions of 3 minutes (around 60 trials) interspersed with a 30-second break were performed. The entire experiment lasted approximately 15 minutes.

**Figure 1.**
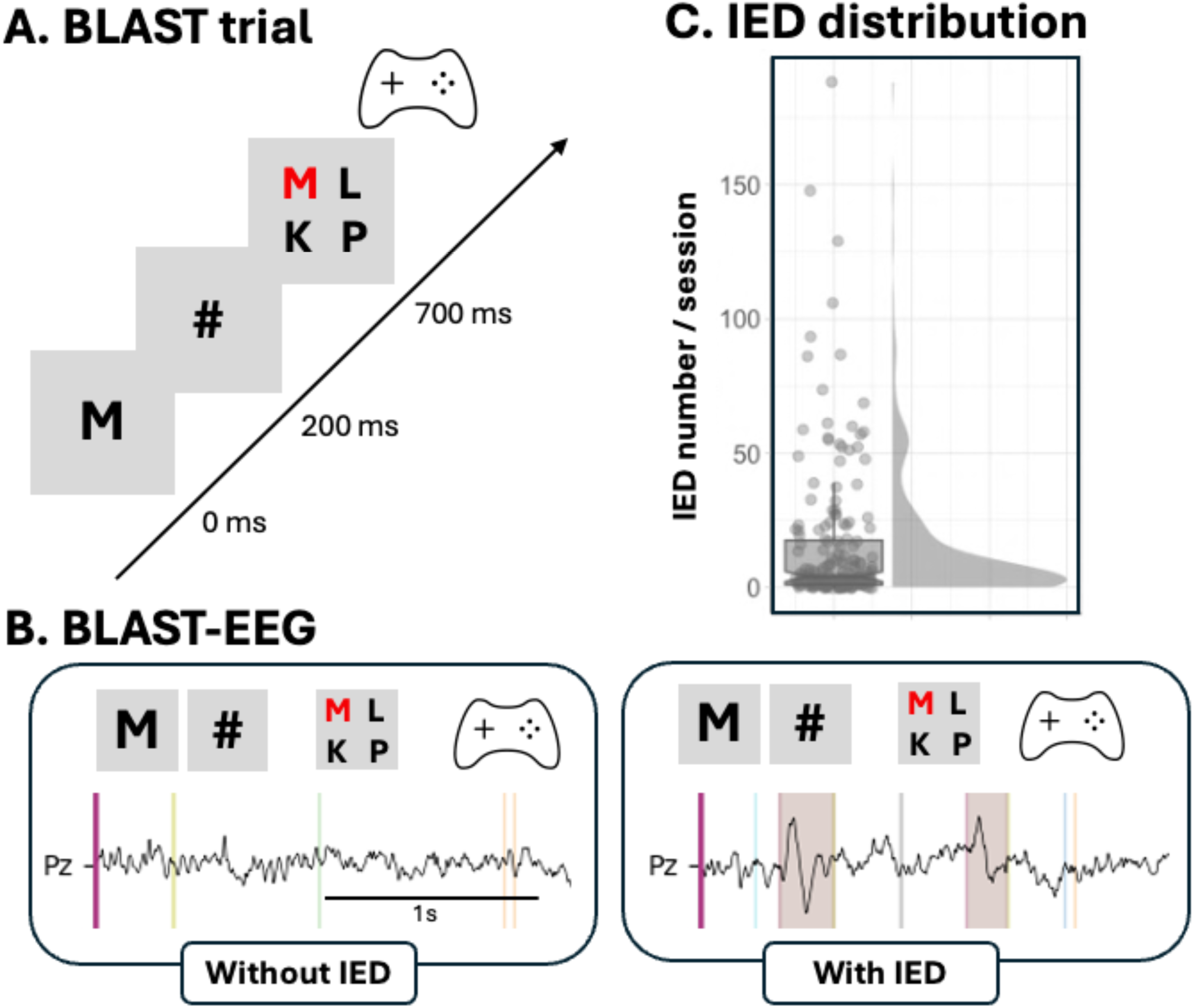
BLAST-EEG paradigm. **A.** Sequential display of a BLAST trial: a letter appears on the screen, followed by a mask, then by an array of 4 letters. Children had to detect the presence (or absence) of a red letter in the array. Answers were given manually using a joystick (yes with the non-dominant hand; no with the dominant hand). The total duration of a session was 3 minutes, with approximately 60 trials to complete. Behavioral and electrophysiological measures were collected. **B.** EEG recordings synchronized with BLAST. Example showing the Pz channel during a trial without (left) and with (right) IED (highlighted). **C.** Distribution of the number of IED in children with at least one IED. Each point represents the number of IED per session (3 sessions per subject). The central line of the boxplot corresponds to the median number of IED per session, the upper and lower parts correspond to the first and third quartiles. Data distribution is represented by density plot (R-ggplot2).

Each trial involves a series of simple cognitive processes, all dependent on the subject’s attentional resources: visual target detection, response selection, and motor transformation. Each trial lasts 2 seconds to measure attention abilities with an excellent temporal resolution. Attention abilities were measured using reaction time (RT) in ms (from 4 letters presentation to motor response), percentage of errors (false-alarms and omissions), as well as BLAST-Intensity and BLAST-Stability, which were created to capture the dynamic of attention over the task. BLAST-Intensity quantifies the ability to produce long series of fast and accurate responses. BLAST-Stability quantifies the ability to produce long series of correct responses with a consistent RT, independently of speed. BLAST-Stability relies on the variation of RT without taking speed into account, focusing on regularity. For both measures (one value per BLAST session), a higher score corresponds to a better stability of attention (i.e. fewer momentary lapses of attention). The full details of computation are provided in the Supplementary materials, Figure S1.

#### 2.2.2. EEG acquisition and processing

EEG activity was recorded from 19 scalp sites (Fp1, Fp2, Fz, F3, F4, F7, F8, Cz, C3, C4, T3, T4, T5, T6, Pz, P3, P4, O1, O2) positioned according to the international 10-20 system, with a mastoid reference, at a sample rate of 1024 Hz using Micromed^®^ Système+ software synchronized with the BLAST acquisition computer. Impedances were kept below 10 KΩ. After recording, an experienced epileptologist (J.D.B) visually identified IED from all channels by determining their start and end points. IED comprised single interictal spikes, polyspikes, sharp waves, or bursts of epileptiform activity. Then, preprocessing was performed using Python (version 3.11.7) including average re-referencing, data filtering with a low-pass at 70 Hz, a high-pass at 1 Hz, and a 50 Hz notch filter when needed, resampling to 256 Hz, and visual rejection of bad channels and segments (*e.g.* containing high amplitude artefacts). A 2-second time window centered on the 4-letter array was defined to create EEG epochs with a baseline correction (from −200 ms to 0 ms), contrasting epochs (BLAST trials) with and without IED (*i.e.* IED occurred within or superimposed on the epoch; **Figure 1**). The amplitude (μV) was extracted every of 0.004s within each epoch.

### 2.3. Statistical analysis

Statistical analyses were performed using R (4.0.4).^19^ Trials with a RT <200 ms and sessions with a percentage of errors >50% were excluded from the statistical analyses, as it reflects unlikely performance. All analyses were only performed on children with epilepsy with at least one IED (**Figure 1**).

#### 2.3.1. Behavioral analyses

Following a descriptive analysis, to model the BLAST scores, generalized linear models (GLM) were implemented via the glm() function from the lme4 package,^20^ specifying the Gamma family with a log-link function. This choice was motivated by the characteristics of BLAST-scores, which were strictly positive and exhibited a pronounced right-skewed distribution. Models were fitted to assess the cumulative effect of IED on BLAST scores: mean RT for correct trials, percentage of errors, BLAST-Stability, BLAST-Intensity. Explanatory variables included IED quantity (number per session), number of ASM (none, mono- or poly-therapy), epilepsy type (focal, generalized), seizure frequency within the last 12 months (none, 1-12, 13-52, 1/day), age at epilepsy onset, session number (1, 2, 3), group (with or without ADHD), and age.^14^ Non-linear effects of IED quantity and age were modeled using second-degree polynomials. Main effects and interactions of interest (session*group, session*age, group*age) were considered using likelihood ratio tests. Post-hoc pairwise comparisons between levels of significant categorical effects have been assessed using the emmeans package.^21^ Estimated marginal means (EMMs) were computed to account for the structure of the fitted model, allowing for marginalization over other factors and covariates. Pairwise contrasts between EMMs were tested, with multiple comparisons corrected using the false discovery rate.

To examine the transient effect of IED on RT for correct trials, a generalized linear mixed model (GLMM) with a Gamma distribution was fitted via the mixed() function from the afex package, including subject as random effect. Fixed effect comprised IED (with *vs* without), session, group and age along with interactions of interest (session*age, group*age, group*IED, age*IED), and were tested via likelihood ratio tests. Visualizations are inspired from Allen *et al*.^22^ Finally, to obtain robust estimates of the effect of IED on RT and address the imbalance between the number of trials with and without IED, a bootstrapped linear regression was performed. For each child with epilepsy, trials without IED were randomly down sampled to match the number of trials with IED. The balanced datasets were resampled with replacement across 10,000 iterations, preserving the within-subject structure in each iteration. A linear model was fitted to each bootstrapped sample using a stepwise regression method with backward elimination. In each model, the intercept represented the mean RT for trials without IED, and the slope captured the RT difference between IED conditions. Distributions of these coefficients across bootstrap samples were summarized to obtain point estimates and 95% confidence intervals (CI).

#### 2.3.2. Electrophysiological analyses

To characterize the temporal dynamics of ERP amplitude, a generalized additive mixed model (GAMM) was employed, as implemented in the mgcv package.^23^ GAMMs extend generalized linear models by allowing non-linear relationships between variables, thereby providing a flexible regression framework. As opposed to traditional ERP analyses, which assume independence across time points, temporal autocorrelation is accounted for, and slight changes in signal amplitude over time are accommodated without imposing rigid *a priori* assumptions about waveform shape. The issue of multiple comparisons is addressed implicitly through penalized smoothing, therefore controlling the effective degree of freedom. Furthermore, GAMMs handle individual variability and trial imbalance (i.e. between IED conditions) by incorporating random smooths (*e.g.* by-subject or by-trial), enabling to capture both group-level trends and individual-level deviations.^23,24^ The GAMM was fitted to the EEG data from −200 to 800 ms around the 4-letter array, with ERP amplitude (μV) averaged across parietal electrodes (Pz, P3, and P4) as the outcome. The main predictor was IED presence (with or without). Non-linear effects of time were modeled separately for each IED condition using interaction smooths with cubic regression splines (k = 20). Individual deviations from the group-level were captured using random smooth of time by subject. To accommodate outliers and deviations from normality, the model used a scaled t-distribution with an identity link. Temporal autocorrelation was addressed using a first-order autoregressive structure (rho = 0.98). Model selection was based on smoothing parameters and overall complexity using itsadug package.^25^ To assess the robustness of the observed effect and address the imbalance between the number of trials with and without IED, a permutation procedure was applied: in each of the 100 iteration, IED labels were randomly permuted, the model was refitted, and the predicted IED effect recalculated. The resulting null distribution under the assumption of no effect of IED was used to define 95% CI. Observed effects falling outside this range were considered statistically significant. For interested readers, supplementary material contains the averaged ERPs (Figure S2).

## 3. Results

### 3.1. Descriptive analysis

Among the 119 included children with epilepsy, 51% presented at least one IED during the BLAST and were selected. The median IED number per session was 15 (range: 0-188) with a median duration of 1.05 s (range: 0.13-22.3). The analyzed cohort was composed of 61 children with epilepsy (56% of girls), the mean age was 9.3 years old (range: 5.9-17.9). Half of the cohort (49%) had a comorbid ADHD. The median age at epilepsy onset was 74 months (range: 5-189) and 54% had a focal epilepsy. Regarding seizure frequency, 28% of children reported no seizure at all, 35% reported 1 to 12 seizures over the last 12 months, 18% 13 to 52 seizures, and 18% at least one seizure per day. Regarding ASM, 33% of children had no medication, 36% had a monotherapy, and 31% a polytherapy. Syndromes are detailed in Supplementary Material, Table S1.

### 3.2. Behavioral analyses

The final fitted models are reported in **Table 1**. Only significant effects (main and interactions) of epilepsy-related features on BLAST scores are presented below.

**Table 1.** Fitted models parameters.

| Models | BLAST measure | IED measure | Epilepsy-related variables | Confounders | Random | R2 |
| --- | --- | --- | --- | --- | --- | --- |
| *Cumulative effect* | Mean RT for correct trials | Quantity | ASM, epilepsy type, seizure frequency, age of onset | Group*age, session | - | 0.716 |
|  | Error percentage | Quantity | ASM, epilepsy type, age of onset | Group*age, session | - | 0.427 |
|  | BLAST-Stability | Quantity | ASM, epilepsy type, seizure frequency, age of onset | Group, age, session | - | 0.345 |
|  | BLAST-Intensity | Quantity | ASM, epilepsy type, seizure frequency, age of onset | Group, age, session | - | 0.259 |
| *Transient effect* | RT for correct trials | Trials with vs without | - | Group, age, session | Subject | 0.540 |

Interactions are reported with a star \* (column 5), R2 stands for the Nagelkerke's R2 of the model for the cumulative effects, and for marginal R2 of the model for the transient effect (column 7). Abbreviations: ASM: antiseizure medication; IED: interictal epileptiform discharges; RT: reaction time.

#### 3.2.1. Cumulative effect of IED

**BLAST-RT.** The mean RT was impacted by the IED quantity (LR Chisq = 15.02, *p* = 0.001): a higher IED quantity was associated with longer RT (**Figure 2A**). The mean RT was also impacted by the age at epilepsy onset (LR Chisq = 5.39, *p* = 0.02) and by the number of ASM (LR Chisq = 18.27, *p* = 0.0001; Supplementary materials, Figure S3 and S4). A younger age at epilepsy onset, was associated with longer RT. The mean RT was higher in children with epilepsy treated by polytherapy compared to monotherapy (estimate = 0.127, t = 2.62, *p^FDR^* = 0.01) or no ASM (estimate = 0.166, t = 3.81, *p^FDR^* = 0.001).

**Figure 2.**
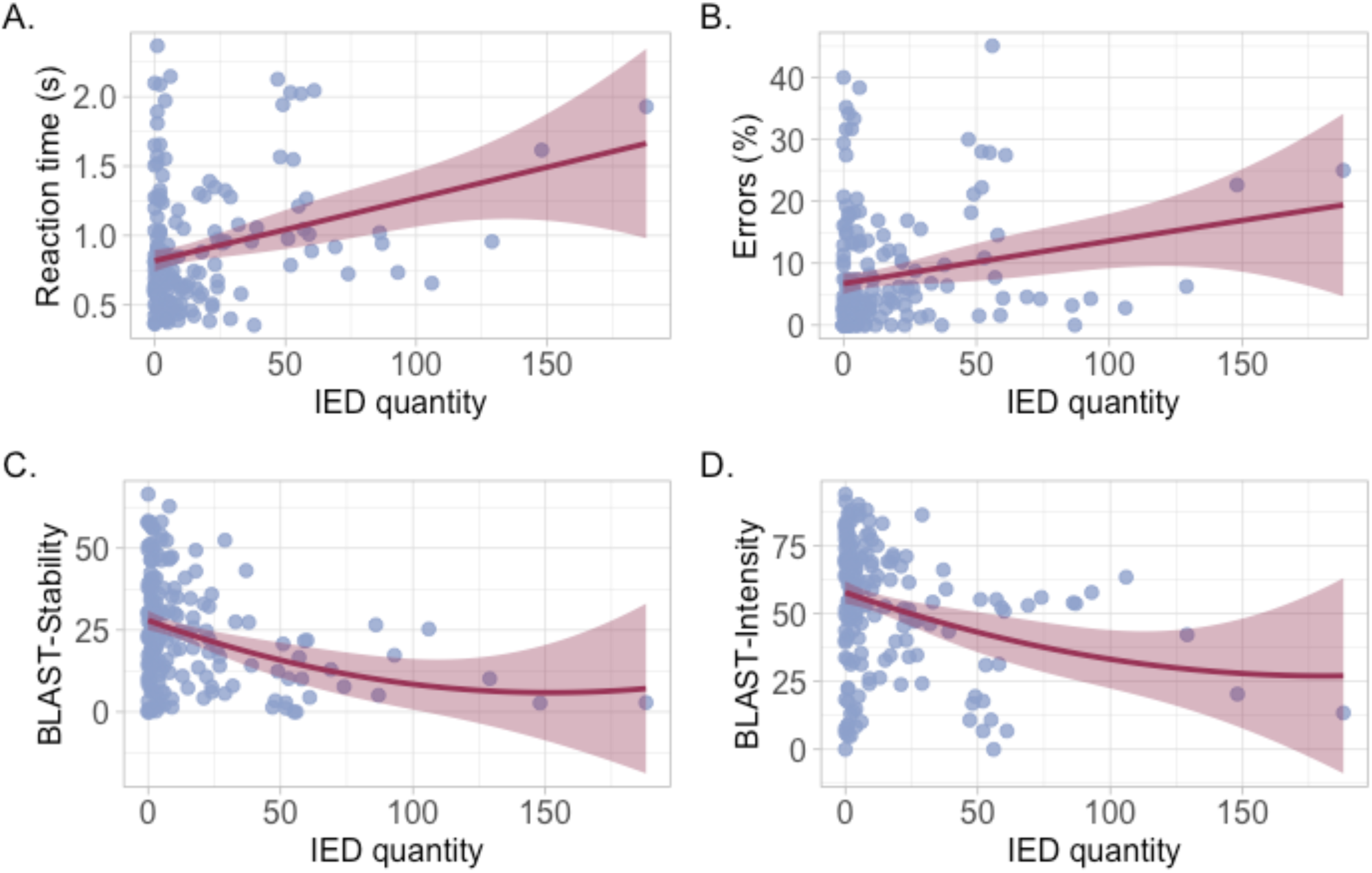
Associations between BLAST-scores and IED quantity. Each point represents (A) the mean reaction time in seconds, (B) error percentage, (C) BLAST-Stability, and (D) BLAST-Intensity per session according to the number of IED. The line represents the linear fit and the light-red band stands for the 95% confidence interval (R-ggplot2).

##### BLAST-Errors

The error percentage was impacted by the IED quantity (LR Chisq = 7.82, *p* = 0.02): a higher IED quantity was associated with a higher error percentage (**Figure 2B**). The error percentage was also impacted by the number of ASM (LR Chisq = 15.54, *p* = 0.001): the error percentage was higher in children with epilepsy treated by polytherapy compared to monotherapy (estimate = 4.35, t = 2.89, *p^FDR^* = 0.01), or in children with epilepsy without ASM compared to monotherapy (estimate = −2.134, t = −2.38, *p^FDR^*= 0.03; Supplementary materials, Figure S4).

##### BLAST-Stability

BLAST-Stability was impacted by IED quantity (LR Chisq = 15.42, *p* = 0.001): a higher IED quantity was associated with lower BLAST-Stability (**Figure 2C**). BLAST-Stability was also impacted by the age at epilepsy onset (LR Chisq = 8.04, *p* = 0.02) and number of ASM (LR Chisq = 20.20, *p* < 0.0001; Supplementary materials, Figure S3 and S4). A younger age at epilepsy onset was associated with a lower BLAST-Stability. The BLAST-Stability was lower in children with epilepsy treated by polytherapy compared to monotherapy (estimate = −12.58, t = −4.46, *p^FDR^* < 0.0001), or no ASM (estimate = −6.15, t = −2.77, *p^FDR^* = 0.01), and higher in children with epilepsy treated by monotherapy compared to no ASM (estimate = 6.44, t = 2.21, *p^FDR^*= 0.03).

##### BLAST-Intensity

BLAST-Intensity was impacted by IED quantity (LR Chisq = 11.38, *p* = 0.003): a higher IED quantity was associated with a lower BLAST-Intensity (**Figure 2D**). BLAST-Intensity was also impacted by the age at epilepsy onset (LR Chisq = 7.36, *p* = 0.03) and the number of ASM (LR Chisq = 15.79, *p* < 0.001; Supplementary materials, Figure S3 and S4). A younger age at epilepsy onset was associated with a lower BLAST-Intensity. The BLAST-Intensity was lower in children with epilepsy treated by polytherapy compared to monotherapy (estimate = −18.40, t = −4.18, *p^FDR^* = 0.0001) or no ASM (estimate = −13.26, t = - 3.16, *p^FDR^* = 0.003).

#### 3.2.2. Transient effect of IED

A significant effect of IED on RT was found (Chisq = 39.95, *p* < 0.001): the RT was significantly increased in trials with IED compared to those without IED (**Figure 3**). Based on 10,000 bootstrap samples, the mean RT for the condition without IED was estimated at 876 ms (95% CI [853-898 ms]). The difference in RT between the 2 conditions was estimated at 79 ms (95% CI [57 ms-102 ms]), suggesting a statistically significant increase in RT in the condition with IED compared to without (Supplementary materials, Figure S5). This effect was consistent across different bootstrap samples.

**Figure 3.**
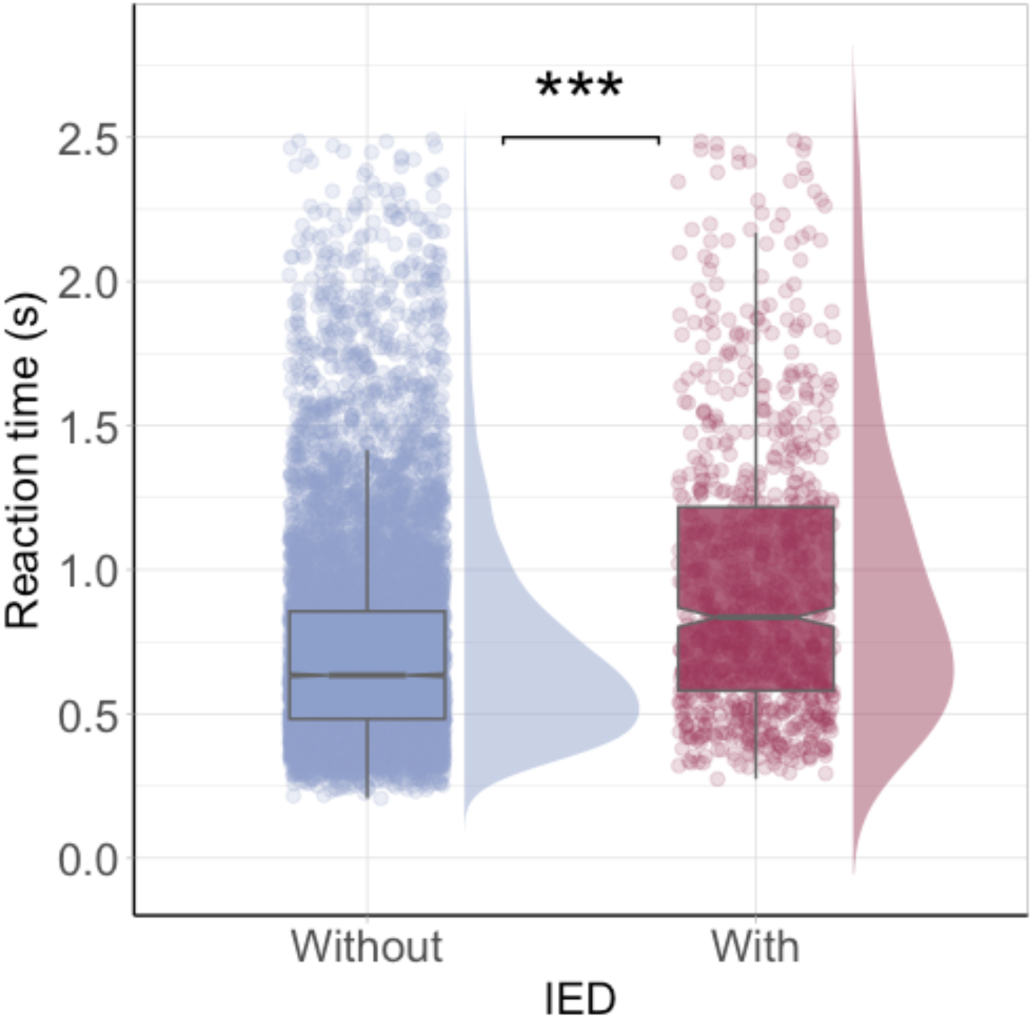
Reaction time for trials without and with IED. Each point represents the RT for each trial without (blue) and with (red) IED. The central line of boxplots corresponds to the median RT; the upper and lower parts correspond to the interquartile range. Data distribution for each condition is represented by density plot. Difference between groups is represented by stars: ***: p ≤ 0.001 (R-ggplot2).

### 3.3. Electrophysiological analyses

The GAMM accounted for 3.49% of the variance in ERP amplitude measured from parietal electrodes, with an explained deviance of 3.46%. This level of explanation reflected significant patterns in the data and successfully captured individual variability in amplitude responses (**Table 2**). A significant main effect of IED was observed: the parietal ERP amplitude was significantly lower in trials with IED compared to those without, and this effect was time dependent. The permutation analysis confirmed the robustness of this finding, showing that the observed difference in amplitude over time exceeded random expectations. Significant time intervals include the period around 300 ms (**Figure 4** for all significant time intervals).

**Figure 4.**
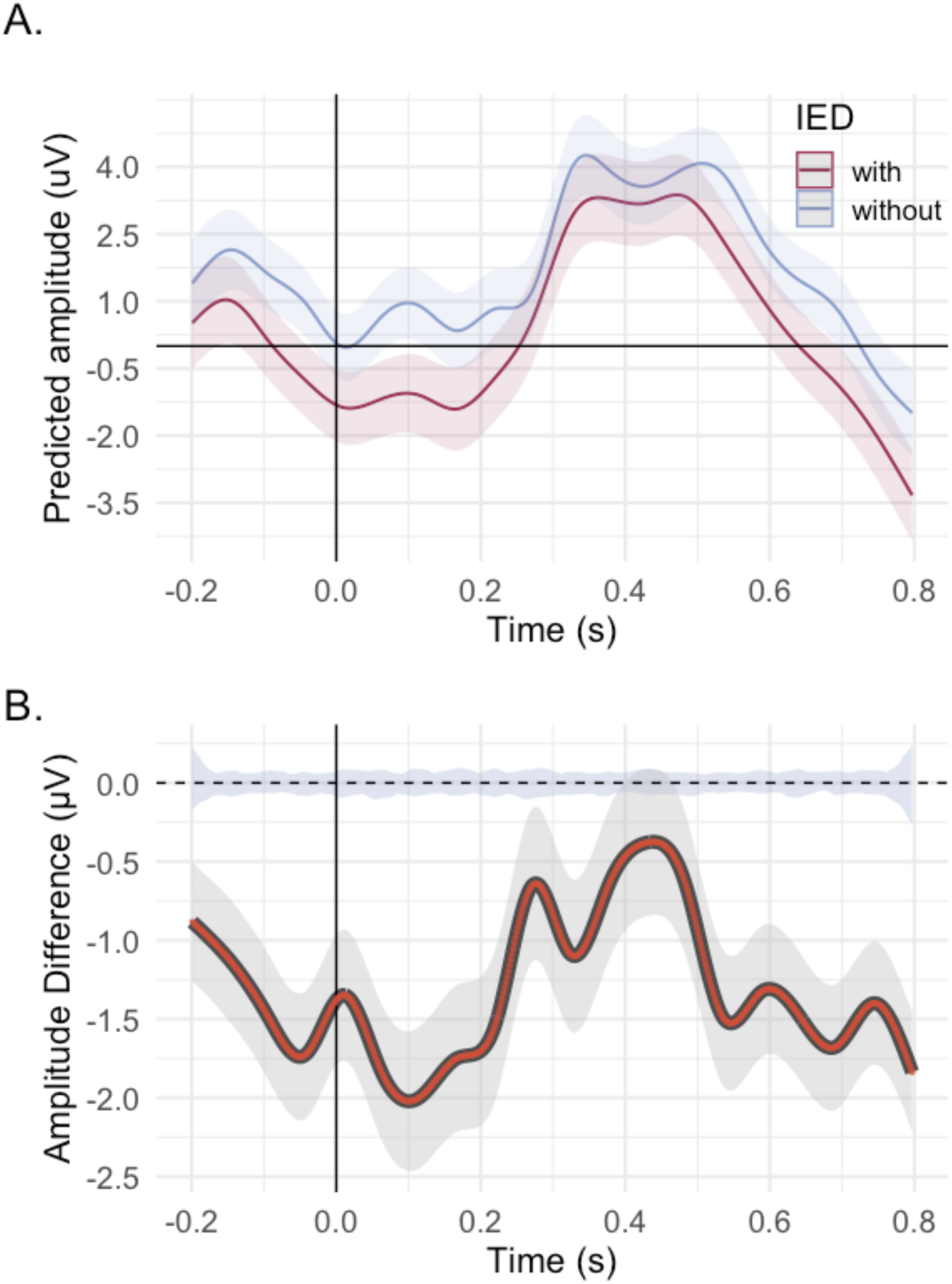
Fitted ERP in parietal electrodes. **A.** Non-linear smooth estimates of EEG amplitude (in microvolts) over time for trials with (red) and without (blue) interictal epileptiform discharges (IED) in combined parietal electrodes (Pz, P3, P4). The shaded bands represent 95% CI. The x-axis represents time in seconds (s), with 0 marking the 4 letters presentation (R-ggemmeans). **B.** Amplitude difference between IED conditions over time. The black curve represents the estimated amplitude difference (with vs without). Negative values indicate greater amplitude in the “without IED” condition. The model includes subject-level random effects, and the curve was smoothed over 600 time points. The shaded blue area represents the 95% CI of the permutation-based null distribution. The red dots superimposed on the difference smooth represents time points where the amplitude difference is statistically significant (i.e. falling outside the shaded null interval).

**Table 2.** Parametric and smooth term estimates from the Generalized Additive Model.

| Parametric terms<br>(linear effects) | Estimate | Standard error | t-value | p-value |
| --- | --- | --- | --- | --- |
| Intercept | 0.373 | 0.316 | 1.180 | 0.238 |
| IED | 1.315 | 0.195 | 6.745 | <0.001 |
| Smooth terms<br>(nonlinear effects over time) | Estimated df | Reference df | F | p-value |
| Time effect for IED with<br>*s(time): IED with* | 16.28 | 19 | 18.91 | <0.001 |
| Time effect for IED without<br>*s(time): IED without* | 17.38 | 19 | 43.09 | <0.001 |
| Subject-specific variation<br>*s(time, patient)* | 973.45 | 999 | 90.22 | <0.001 |

Abbreviations: IED: interictal epileptiform discharges; df: degree of freedom.

## 4. Discussion

### IED induce fluctuations of attention

Overall, the present findings emphasized the deleterious impact of IED on attention; the data suggested that IED act as internal micro-distractors as they induce both cumulative and transient attentional fluctuations. The increased quantity of IED was associated with an increase in RT and errors, and a decrease in BLAST-Stability and BLAST-Intensity. Cumulative effects on cognition have already been highlighted when IED quantity on EEG exceeds a certain threshold (*e.g.* >1 or 10%) or in specific epileptic syndromes with abundant diurnal or nocturnal IED.^5,26–30^ However, in previous studies, challenges mainly lied in distinguishing IED’s effects from those of other epilepsy-related features.^31^ The present results provided evidence that the impact of IED is independent from other clinical factors comprising: seizure frequency, age at epilepsy onset, epilepsy type, number of ASM, or presence of a comorbid ADHD. Among these features, a younger age at seizures onset or a higher number of ASM (starting from two ASM or more), was associated with a worst attention performance, as already reported.^5,30,32^ It was further observed that IED have more subtle impact, responsible for momentary lapses of attention; the trials with IED have been characterized by a mean increase in RT greater than 75 ms, which, given the simplicity of the task, can be considered as a meaningful delay. In parallel, a significant and sustained down-modulation of the parietal ERP amplitude was observed in these trials, extending over a long-time window that include the classical P300 period. These findings suggest that IED do not only affect general neural excitability but also alter stimulus-driven cognitive processes, such as attention allocation and stimulus evaluation. These results align with the well-known concept of transient cognitive impairment (TCI),^4^ described as the temporary incapacity to effectively complete a task during IED occurrence. Since IED impacts are not easy to demonstrate due to multiple confounding factors such as intersubject variability or underlying diseases,^12^ TCI can only highlighted through suitable testing. For instance, the study of auditory evoked responses or more recent studies on intracranial EEG using trial-by-trial analysis contributed to highlight a deleterious impact of IED on various cognitive functions (*e.g.* flexibility, word-finding, memory encoding).^12,13,33,34^ As traditional attentional tests lack the sensitivity to demonstrate an altered response concomitant to IED, the BLASTEEG paradigm meets important clinical needs. Ergonomic, easy-to-use, and well-accepted by children, this protocol aligns with the time constraints of real-life testing and could provide help in guiding therapeutic strategies. Considering the high frequency of IED in routine EEG, their impacts on day-to-day activities such as reading or driving must be assessed, especially in developing brain due to potential long-term repercussions.

### IED could disrupt large-scale synchronization/desynchronization processes

Regarding the mechanisms underlying the deleterious effects of IED on attention stability, several could be suggested. First, although cognitive deficits have been linked to the specific brain regions in which focal IED occur,^4,6–8,33^ attention impairment was observed herein without considering IED topography, consistent with a recent data from the literature.^13^ However, given the extensive coverage of attention networks,^35,36^ their disruption by IED is a highly probable outcome. Second, IED may trigger large-scale neurophysiological changes,^37^ including the spread of aberrant rhythmic activity and disruptions in functional networks organization.^37–41^ This aligns with EEG-fMRI studies that found that IED can alter activation-deactivation patterns, sometimes beyond the epileptic focus.^8,31,42–44^ Performance on BLAST relies on a dynamic interplay between the activation of the dorsal attention network and the deactivation of default-mode network.^14^ IED may disturb this balance, mirroring the patterns observed during momentary lapses of attention.^45^ Future intracranial EEG studies are needed to clarify how IED disturb the gamma-band activity within the BLAST-related networks. In summary, IED may act as transient but impactful disruptions of functional networks, thereby involved in attention impairments.

### Limitations and perspectives

Sheybani *et al.,* recently reported that local slow waves during wakefulness constitute a homeostatic process that responds to changes in network excitability and reduces aberrant activity associated with IED at the expense of TCI.^46^ While this study excluded 1-second post-IED slow waves, previous evidence indicated that slow-waves following IED were responsible for the bulk of the cognitive impairment.^47^ The type of IED was not taken into account herein, allowing the possibility that each, or both components (including the wave), may have been involved in the observed impairment. An additional question pertains to the relationship between IED and vigilance. IED have been previously found to occur more frequently during periods of drowsiness or inattention, and less frequently during the active engagement into a cognitive task.^4,44,48^ Therefore, the role of vigilance variation in the modulation of both IED occurrence and attention performance must be further investigated. Finally, the present study has limitations: (i) the simplicity of the task, which may have led to an underestimation of IED impact on attention performance, especially in older children; and (ii) the heterogeneity of syndromes. While previous studies have suggested that changes in network levels related to IED are generalizable across patient groups and clinical features,^40^ investigating the relationship between IED and attention disorders by discriminating etiologies could be the subject of future research.

## 5. Conclusions

The present study using BLAST-EEG suggests that IED are responsible for attentional fluctuations; a clear attention alteration concomitant with IED occurrence was identified by examining moment-to-moment behavioral responses and ERP in a large, unselected cohort of children with epilepsy. IED impacts were discriminated from those of other epilepsy-related features. This work provides evidence that IED induce subtle attentional deficits and could be considered as biomarkers of abnormal brain function.

## Data availability

This work is based on clinical data that are not available due to ethical and privacy restrictions: consent forms do not allow the use of data by other research teams. Data may be available upon reasonable request from the corresponding author and after additional and documented consent of patients and caregivers involved in this study.

## Code availability

Code for electrophysiological and statistical analyses are available at https://github.com/mthieux/IED_BLASTEEG

## Acknowledgements

This study is part of M.T. postdoctoral project funded by the Université Claude Bernard Lyon 1 (UCBL; AAP ETOILES 2023). We thank Shanez Haouari for her help in manuscript preparation.

## Author contributions

Designed the study: R.B., J.D.B, V.H., J.J., P.K., J-P.L., M.P-B., M.T., L.V.

Data acquisition: J.D.B., V.H., M.T.

Analyzed the data: R.B., L.M., M.T.

Wrote first draft of the manuscript: M.T.

All authors reviewed on subsequent drafts.

## Competing interests

The authors declare no competing interests.

## Notes

### Competing Interest Statement

The authors have declared no competing interest.

### Funding Statement

This study is part of M.T. postdoctoral project funded by the University Claude Bernard Lyon 1 (UCBL; AAP ETOILES 2023).

### Author Declarations

The study protocol was approved by the French Comite de Protection des Personnes (Sud Est V) (COGNIT-AIC-38RC14.374) and recorded on ClinicalTrials.gov (ID: NCT03094793).

