## Supplementary materials for "Interictal epileptiform discharges are involved in momentary lapses of attention in children with epilepsy"

**Table S1.** Clinical syndromes.

| | **Children with epilepsy**<br>N = 61 |
|---|---|
| Focal epilepsy | |
| Symptomatic / cryptogenic | 21% (13) |
| Self-limited with centrotemporal spikes | 18% (11) |
| Self-limited with autonomic seizures | 11% (7) |
| Unknown causes | 3% (2) |
| Generalized epilepsy | |
| Childhood absence | 10% (6) |
| Juvenile absence | 2% (1) |
| Juvenile myoclonia | 2% (1) |
| Other idiopathic | 18% (11) |
| Symptomatic | 2% (1) |
| Unknown causes | 5% (3) |
| Eyelid myoclonia | 8% (5) |

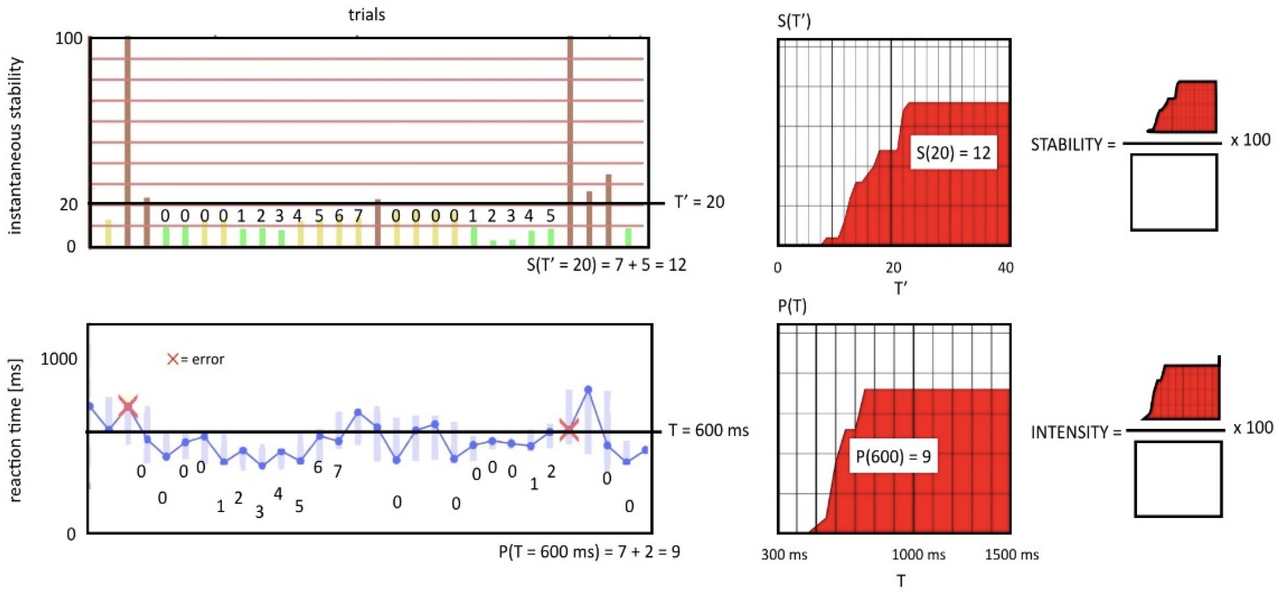


**Figure S1. Scoring systems for BLAST-Intensity and BLAST-Stability**. BLAST-Stability (top) and BLAST-Intensity (bottom) are computed from graphs S(T') and P(T), from the ratio of the area under the curve (red) divided by the theoretical maximum area (white), i.e. the maximum score. **BLAST-Intensity** derives from the assumption that highly focused individuals tend to respond fast and with few errors. A performance graph showing RT for all the trials should display long series of hits below a given time limit T (horizontal line), even when T is low. If we cumulate the length of all successful series below T (i.e. with a RT consistently faster than T), we reach a measure P(T) which should be high for highly focused individuals even when T is small. To penalize errors and to be consistent with the adaptive designs which required participants to generate successful series of five trials, we computed a measure P(T), which included only series longer than five consecutive wins, cumulating N-5 points for every such series (where N is the length of that series). P(T) is the number of points for time limit T, it is computed for every value of T between 300 ms and 1500 ms. The bottom left graph illustrates the calculation of P(T) from the RT for an example value of T (600 ms: 9 points). The procedure is repeated for all T to generate red plot P(T). BLAST-Intensity is defined by 100*AUC(P)/max(AUC), where AUC is the area under the curve P(T) and max (AUC) its maximal theoretical value. One limitation of BLAST-Intensity is that it is mathematically lower for participants with slower RT. Yet, someone with slow but constant RT, and no errors, is fully "on-task". **BLAST-Stability** emphasizes stability more than speed and was computed from the instantaneous stability of RT r(trial i). The top left graph illustrates the calculation of S(T') from the instantaneous standard deviation s(trial) of the normalized reaction times (r'(trial)) for an example value of T' (20: 12 points). To minimize the effect of the overall speed, we considered the stability of normalized RT (i.e. RT expressed in % of the median RT). The instantaneous stability of attention for trial i as instab(trial i) = std(r'(trial i-1):r'(trial i+1)) is computed over sliding windows of 3 consecutive trials. To penalize errors, instab was set to a maximal value (40) for unsuccessful trials. We devised a scoring system S(T') which accumulates N-5 points for each series of N > 4 winning trials for which instab stays below T' (a procedure repeated for T' values between 0 and a maximum of 40). BLAST-Stability is defined by 100*AUC(S)/max(AUC), where AUC is the area under the curve S(T') and max (AUC) its maximal theoretical value. Figure from Petton *et al.* (2019).[14]

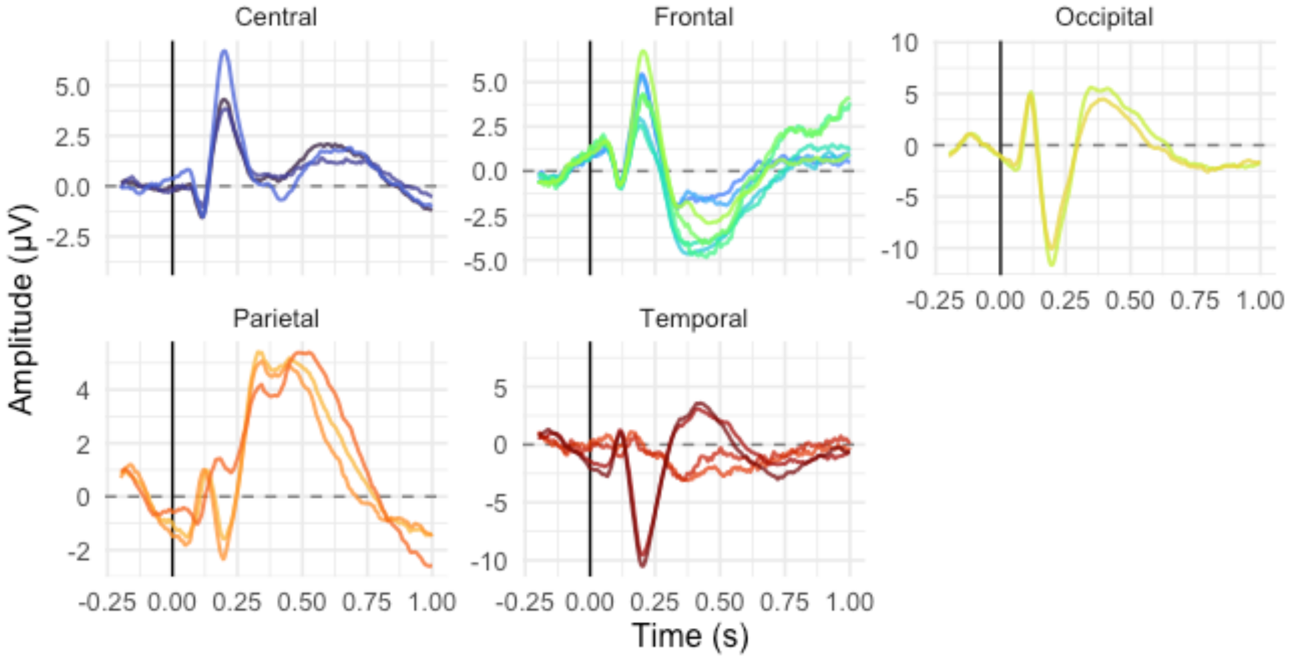


**Figure S2. Grand average EEG signal by channel and region**. Event-related potentials recorded from central (blue), frontal (green), occipital (yellow), parietal (orange), and temporal (red) regions. The x-axis represents time in seconds, with 0 marking the 4-letters presentation. The y-axis shows amplitude in μV averaged for both IED conditions (with, without) (R-ggplot2).

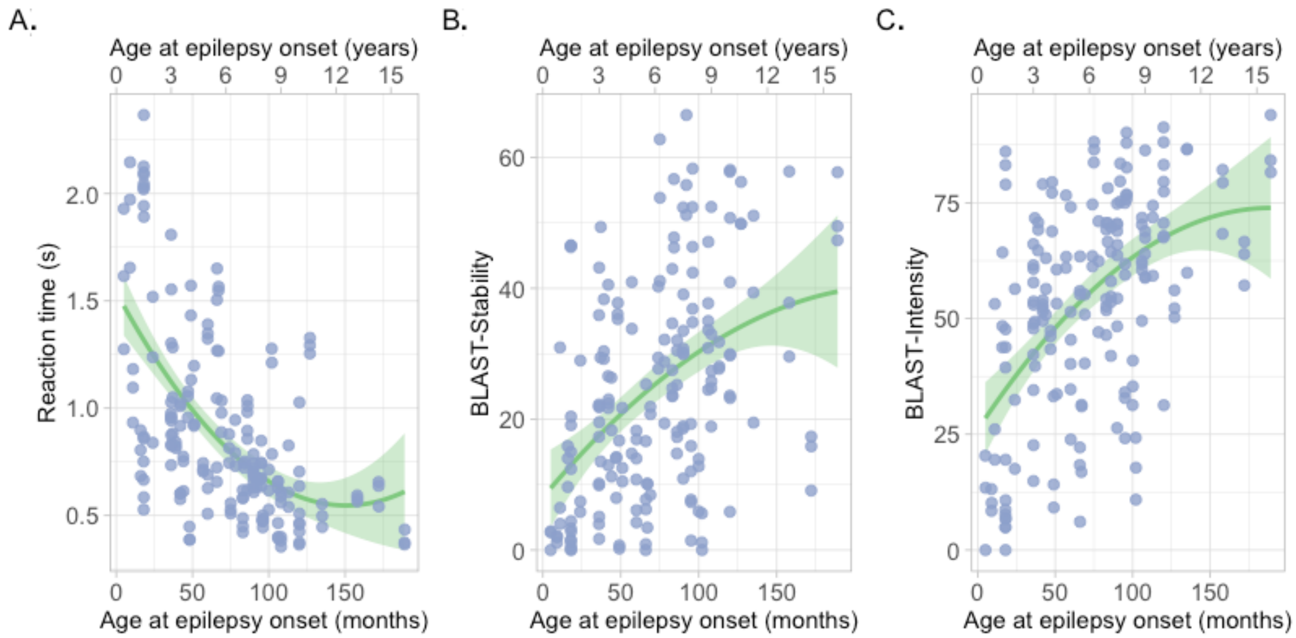


**Figure S3. Associations between BLAST-scores and age at epilepsy onset**. Each point represents (A) the reaction time, (B) BLAST-Stability, and (C) BLAST-Intensity per session according to the age at epilepsy onset (in months). The green line represents the linear fit and the light-green band stands for the 95% CI (R-ggplot2).

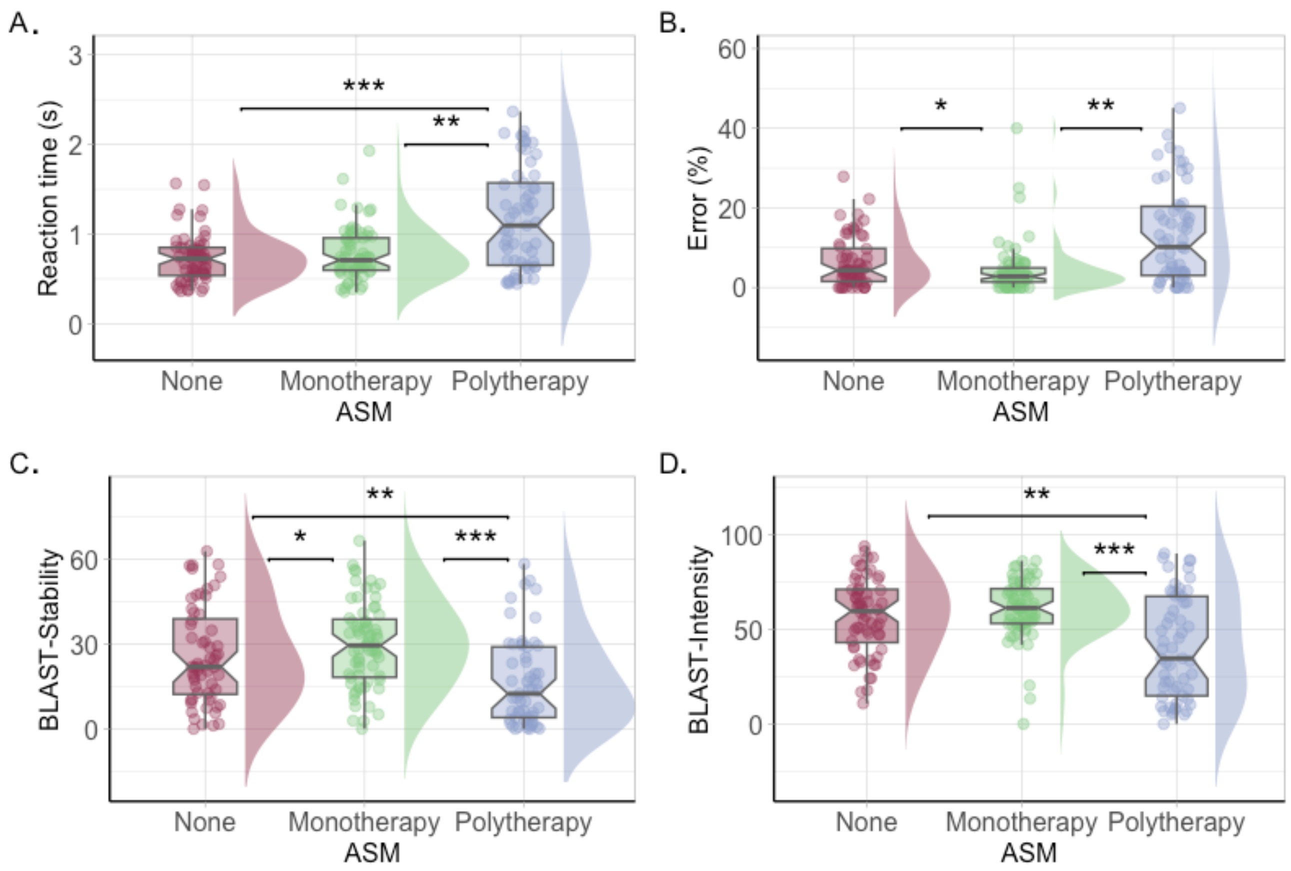


**Figure S4. BLAST-scores according to the number of ASM**. Each point represents (A) the reaction time, (B) percentage of errors, (C) BLAST-Stability, and (D) BLAST-Intensity per session according to the number of ASM (red: none, green: monotherapy, blue: polytherapy). The central line of boxplots corresponds to the median of each score, the upper and lower parts correspond to the first and third quartiles. Data distribution for each condition is represented by density plot. Difference between groups is represented by stars: *: $p \leq 0.05$, **: $p \leq 0.01$, ***: $p \leq 0.001$ (R-ggplot2).

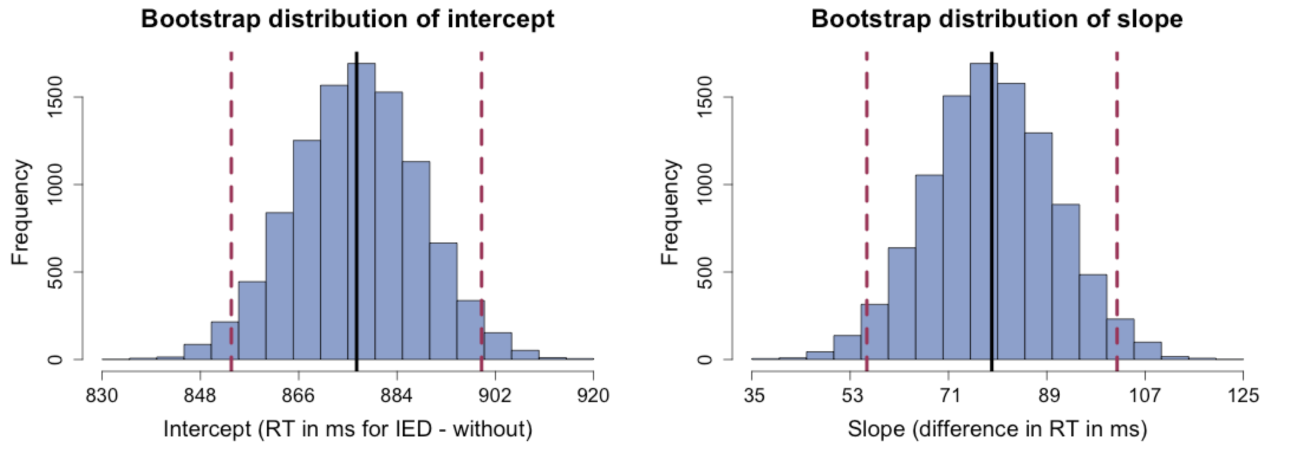


**Figure S5. Bootstrap distributions of reaction time (RT) for trials with and without IED**. Intercept (left) and slope (right) distributions of 10,000 bootstrapped samples based on a linear model assessing the effect of IED (with vs without) on RT, accounting for variability between subjects. The intercept represents the RT (in ms) for trials without IED (baseline condition). The slope represents the difference in RT (in ms) between trials with versus without IED: a positive slope indicates that the "with IED" condition results in longer RTs compared to the "without IED" condition. Black lines represent the means across all samples. Red dotted lines represent 95% CI.
